# “Clinical, Radiological and Therapeutic Characteristics of Patients with COVID-19 in Saudi Arabia”

**DOI:** 10.1101/2020.05.07.20094169

**Authors:** Mohammed Shabrawishi, Manal M Al-Gethamy, Abdallah Y Naser, Maher A Ghazawi, Ghaidaa F Alsharif, Elaf F Obaid, Haitham A Melebari, Dhaffer M Alamri, Ahmad S Brinji, Fawaz H Al Jehani, Wail Almaimani, Rakan A Ekram, Kasim H Alkhatib, Hassan Alwafi

## Abstract

**BACKGROUND:** Coronavirus disease 2019 (COVID-19) is a rapidly spreading global pandemic. The clinical characteristics of COVID-19 has been reported; however, there are limited researches that investigated the clinical characteristics of COVID-19 in the Middle East. The aim of this study is to investigate the clinical, radiological and therapeutic characteristics of patients diagnosed with COVID19 in Saudi Arabia.

**METHODS:** This study is a retrospective single-centre case series study. We extracted data for patients who were admitted to the Al-Noor Specialist hospital with a PCR confirmed SARS-COV-2 between 12th and 31st of March 2020. Descriptive statistics were used to describe patients’ characteristics. Continuous data were reported as mean ± SD. Chi-squared test/Fisher test were used as appropriate to compare proportions for categorical variables.

**RESULTS:** A total of 150 patients were hospitalised for COVID-19 during the study period. The mean age was 46.1 years (SD: 15.3 years). Around 61.0% (n= 90) were males and six patients (3.9%) reported working in the healthcare sector. The most common comorbidities were hypertension (28.8%, n= 42) and diabetes mellitus (26.0%, n= 38). The majority of the patients, 64.4% (n = 96) had a recent contact history with a COVID patient. Regarding the severity of the hospitalised patients, 105 patients (70.0%) were mild, 29 (19.3%) were moderate, and 16 patients (10.7%) were severe or required ICU care. From the 105 mild patients, around 31.3% (n= 47) were asymptomatic.

**CONCLUSION:** This case series provides clinical, radiological and therapeutic characteristics of hospitalised patients with confirmed COVID-19 in Saudi Arabia.

## 1. INTRODUCTION

In early December 2019, a cluster of acute pneumonia of unknown aetiology has been identified in Wuhan, China (1). The pathogen has been identified as a new RNA virus from the betacoronavirus family, and has been named as severe acute respiratory syndrome coronavirus 2 (SARS-CoV-2) (2). The respiratory illness caused by the 2019 novel coronavirus disease (COVID-19) is highly infectious, and therefore, the World Health Organization (WHO) has characterized the diseases a pandemic infection (3). As of April 25, 2020, more than 2,700,000 confirmed cases were reported worldwide, and it has spread from Wuhan to more than 200 countries across the world (4).

The Kingdom of Saudi Arabia (KSA) is the largest country in the Arabian Peninsula and it is located in the South West part of Asia (5). In a historical decision, KSA has suspended Umrah and all religious visits to the country in an attempt to prevent and delay the spread of COVID-19 in KSA. However, in March 2, 2020, Saudi Arabia has confirmed its first case of COVID-19 which was imported from Iran (4). Several other local clusters were identified later with the majority of the cases being linked to recent travel history.

In recent studies, the clinical features and severity of COVID-19 has been described to be similar of other respiratory viruses such as severe acute respiratory syndrome (SARS) and Middle East respiratory syndrome (MERS) (6, 7). Symptoms can range from mild flu-like symptoms to acute respiratory distress syndrome (ARDS) (8). However, the characteristics and the course of the disease in Middle Eastern populations remains unclear. Exploring the clinical characteristics of patients diagnosed with COVID-19 in Saudi Arabia is important knowing that there are many visitors who travel to Saudi Arabia for religious purposes. Beside this, there is a high air traffic for other purposes in this country which was estimated to be around 39 million people in 2018. In 2019, around 7.5 million Muslim entered the holy city of Mecca for Umrah purposes (9). This highlights how crucial is to have a deeper exploration on the characteristics of patients diagnosed with this widespread infection in this region. To address the above knowledge gaps and giving the ongoing spread of COVID-19 in the Middle East, this study aims to describe the clinical, radiological, and therapeutic characteristics of COVID-19 in a selected cohort of patients in Mecca, Saudi Arabia.

## 2. METHODS

### 2.1. Study Design and participants

This was a retrospective single-centre case series study of 150 patients diagnosed with COVID-19. We extracted data for patients who were admitted to Al-Noor Specialist hospital with a polymerase chain reaction (PCR) confirmed SARS-COV-2 between 12^th^ and 31^st^ of March 2020. Al-Noor Specialist hospital in Mecca, Saudi Arabia is a 500-bedder specialist and teaching hospital in the centre of the holy city of Mecca. It delivers tertiary care throughout the Mecca region of Saudi Arabia and its part of the Ministry of Health services (10, 11). All patients enrolled in this study were diagnosed with COVID-19 through real time (RT)-PCR obtained through nasopharyngeal swabs. All data including outcomes, mortality and length of stay were monitored up to 8^th^ April 2020.

### 2.2. Data Collection

Data were extracted from both paper and electronic records using a unique medical record number (MRN) for each patient. All data were reviewed and checked by a medical team including; two medical residents and a consultant Pulmonologist. Extracted data included patients’ demographics, comorbidities, history of recent travel and history of contact with a confirmed COVID19 patient in the past two weeks. In addition, clinical signs, symptoms, radiological findings and pharmacological treatment received were collected. The radiological examinations were interpreted by a certified consultant radiologist who was blinded from the clinical presentation of the patients. The severity assessment of the chest x-ray (CXR) were estimated subjectively. All data were collected at the time of the admission.

### 2.3. Study variables

Data regarding the clinical progression and severity of the disease were reported as the worst classification reached at any point during hospitalisation. We further classified the severity of the disease based on the following criteria; 1) mild disease was defined as patients with upper respiratory tract symptoms (as rhinorrhoea, sore throat, headache, myalgia, body pain, low grad fever and or dry cough) with absent of clinical or radiological finding of pneumonia; 2) moderate disease defined as symptomatic patients with radiological sign of pneumonia; 3) severe disease defined as confirmed COVID-19 pneumonia with any of the following respiratory rate ≥30/min, blood oxygen saturation ≤93% at rest, PaO2/FiO2 ratio <300, lung infiltration >50% of the lung field, and 4) critically sever disease defined as any of the following: respiratory failure required invasive mechanical ventilation, shock or organ failure require admission to the intensive care unit.

### 2.4. Ethics

This study was approved by the institutional ethics board at the Ministry of Health in Saudi Arabia (No. H-02-K-076-0420-286). Patients were informed that their clinical data will be used for clinical or research purposes with keeping all their personal information confidential.

### 2.5. Statistical Analysis

Descriptive statistics were used to describe patients’ demographic characteristics, radiological findings, medications use, and comorbidities. Continuous data were reported as mean ± SD, and categorical data were reported as percentages (frequencies). Independent sample t test was used to compare the mean value for continuous variables. Chi-squared test/Fisher test were used as appropriate to compare proportions for categorical variables. Logistic regression analysis was used to identify predictors of clinical characteristics. A confidence interval of 95% (p < 0.05) was applied to represent the statistical significance of the results and the level of significance was assigned as 5%. SPSS (Statistical Package for the Social Sciences) version 25.0 software (SPSS Inc) was used to perform all statistical analysis.

## 3. RESULTS

### 3.1. Patients clinical characteristics

**Table 1** presents patients’ characteristics at presentation to the hospital. A total of 150 patients were hospitalised for COVID-19. The mean age was 46.1 years (SD: 15.3 years), and ranged between 11 and 87. Around 61.0% (n= 90) were males. Six patients (3.9%) reported working in the healthcare sector. The most common comorbidities were hypertension (28.8%, n= 42) and diabetes mellitus (DM) (26.0%, n= 38). The majority of the patients (56.0%; n= 84) were local resident. Around half of the patients (54.1%, n= 80) reported that they had a contact history with a traveller. In addition, the majority of the patients, 64.4% (n = 96) had a contact history with a COVID-19 patient. Regarding the severity of the hospitalised patients, 105 patients (70.0%) were mild, 29 (19.3%) were moderate, and 16 patients (10.7%) were severe or required ICU care. Of the 105 mild patients, around 31.3% (n= 47) were asymptomatic. Patients with comorbidities were more likely to have a severe outcome compared to other patients (p<0.05). Patients who reported a contact history with a COVID-19 patient were more likely to have mild to moderate severity of the disease (p<0.05). Mild cases were more prevalent among females, while moderate to severe and or critical were prevalent among males (**Figure 1**). For symptomatic patients, the most common symptoms at presentation were fever (49.3%, n= 72), dry cough (48.6%, n= 71), and shortness of breath (19.9%, n= 29) **(Table 2)**. Furthermore, during admission, fever and cough (28%) were the most common symptoms followed by nausea and vomiting (12%). Most of the asymptomatic patients were females (OR: 0.45 [95%CI 0.22 – 0.92]; p=0.027). In addition, patients who reported travel history or a contact with a traveller recently were three times (OR: 3.13 [95%CI 1.52 – 6.45]; p= 0.002) and four times (OR: 4.03 [95%CI 1.84 – 8.81]; p= 0.000) more likely to be asymptomatic, respectively. Besides, patients who have reported a contact with COVID-19 patients were four times at higher risk of being symptomatic (OR: 4.50 [95%CI 1.84 – 10.99]; p= 0.001).

**Figure 1:**
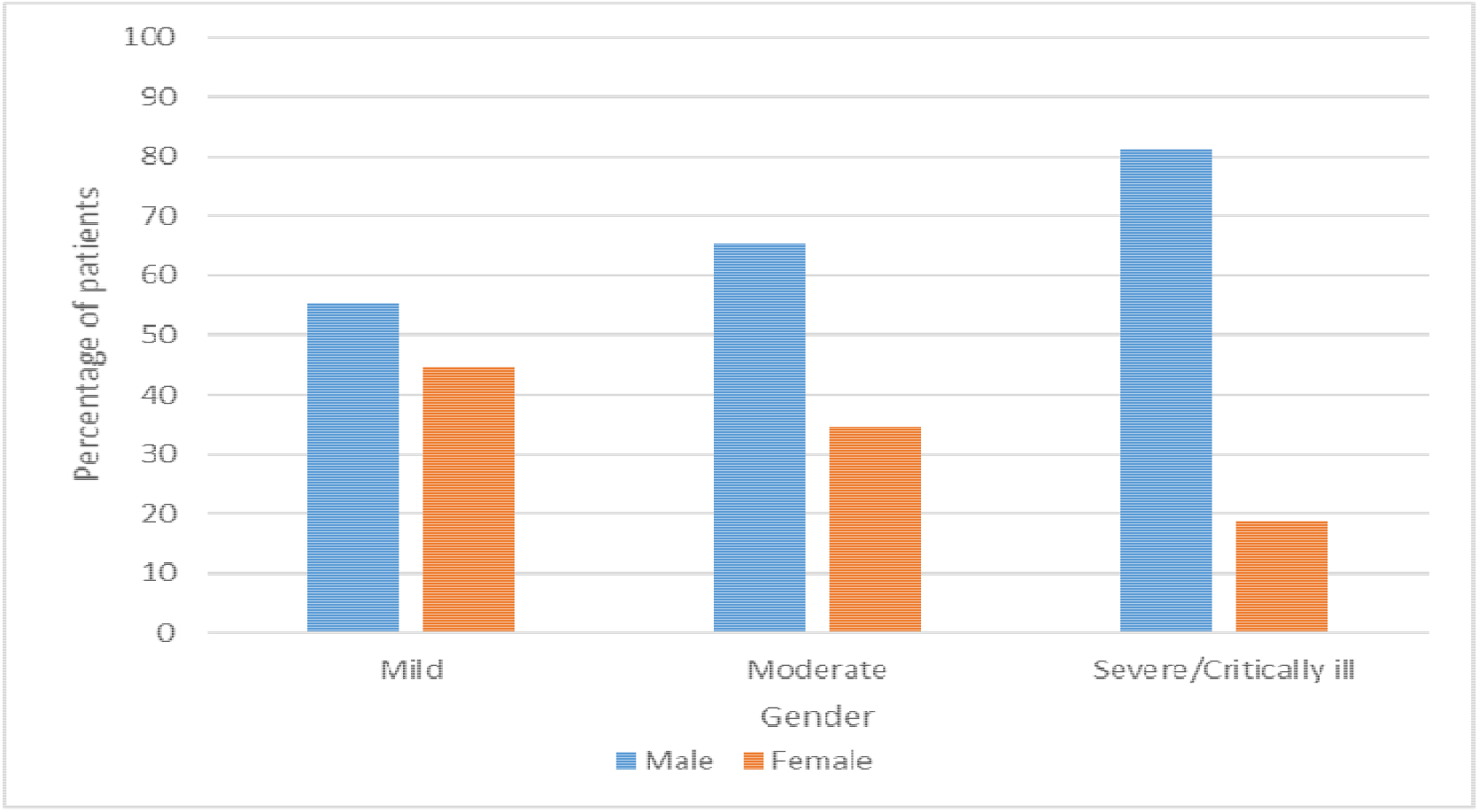
Clinical severity stratified by gender.

**Table 1:**
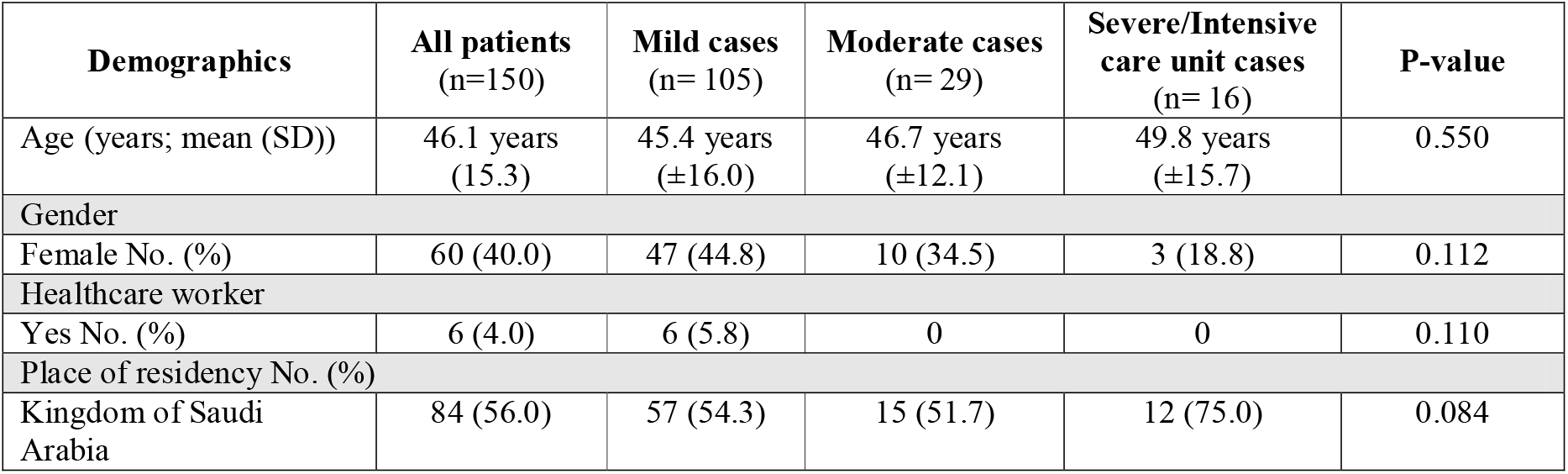
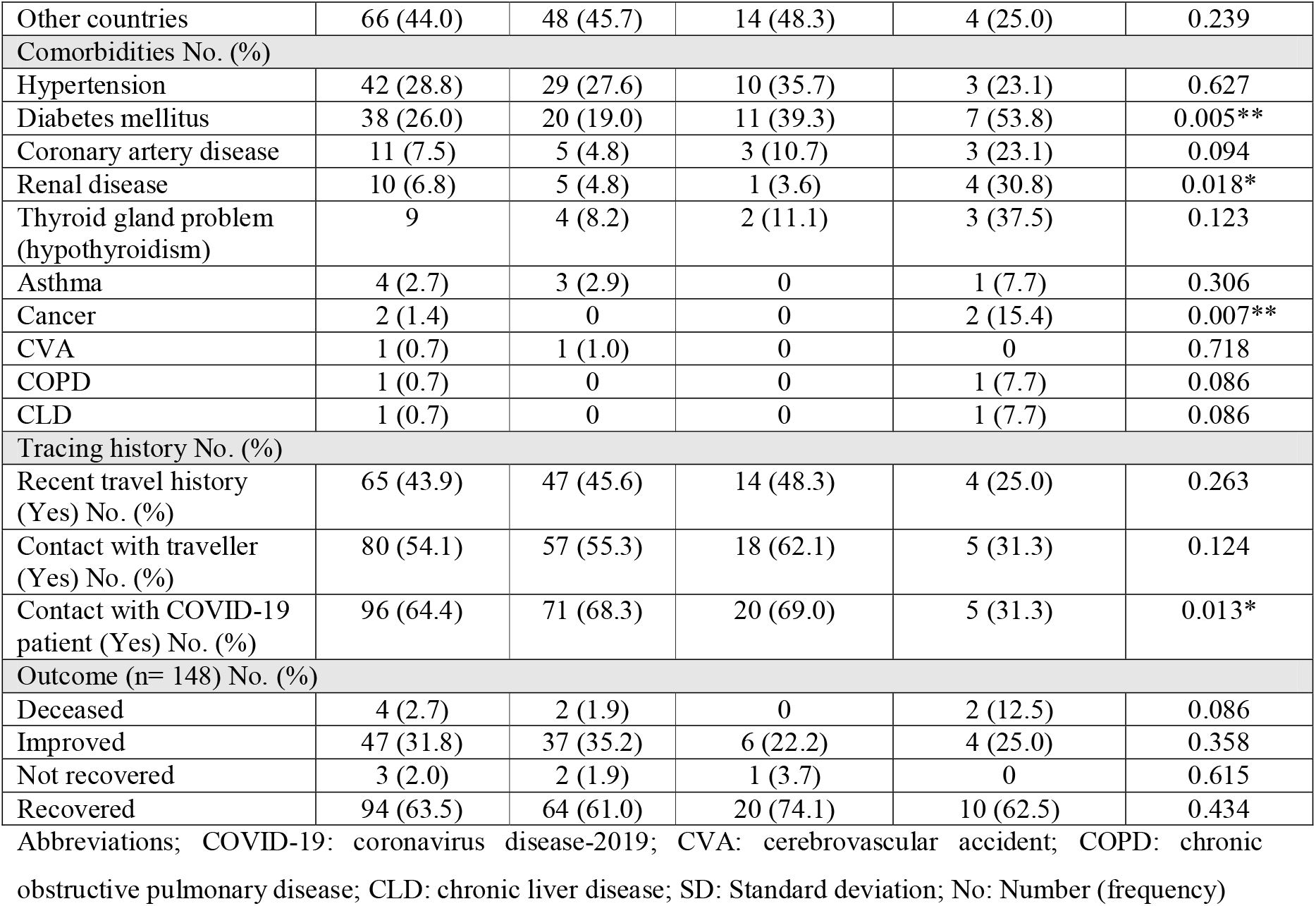
Patients demographic characteristics at presentation.

**Table 2:** Patient signs and symptoms at presentation and during admission.

| Variable | Symptoms |  | P-vale |
| --- | --- | --- | --- |
|  | At presentation No. (%) | During admission No. (%) |  |
| Fever | 72 (49.3) | 28 (19.2) | 0.029* |
| Cough | 71 (48.6) | 28 (19.2) | 0.024* |
| Shortness of breath | 29 (19.9) | 7 (4.8) | 0.000*** |
| Sore throat | 24 (16.4) | 2 (1.4) | 0.269 |
| Runny nose | 9 (6.2) | 0 (0.0) | >0.99 |
| Sputum | 5 (3.4) | 1 (0.7) | 0.034* |
| Headache | 4 (2.7) | 0 (0.0) | >0.99 |
| Myalgia | 4 (2.7) | 1 (0.7) | 0.813 |
| Diarrhea | 2 (1.4) | 5 (3.4) | 0.068 |
| Nausea/vomiting | 1 (0.4) | 12 (8.2) | 0.678 |
| Haemoptysis | 1 (0.4) | 1 (0.7) | 0.887 |
| Fatigue | 1 (0.4) | 1 (0.7) | 0.907 |

\* p<0.05; \*\*p<0.01; \*\*\*p<0.000

### 3.2. Radiological findings

Around half of the patients (49.7%, n= 72) had normal radiological exam at presentation. The severity of the cases was correlated with an increase in the prevalence of GGO at presentation (P=0.002). The predominant pattern of abnormality observed was ground-glass opacification (29.0%, n= 42), peripheral (57.5%, n= 42), and (bilateral (35.3%, n= 35), which was mainly involving the lower lobes (Figure 2). Most of the patients had stable radiological exams on follow up. Around 64.6% (n= 62) showed progression, half of them belongs to more severe group **(Table 3)**.

**Figure 2:**
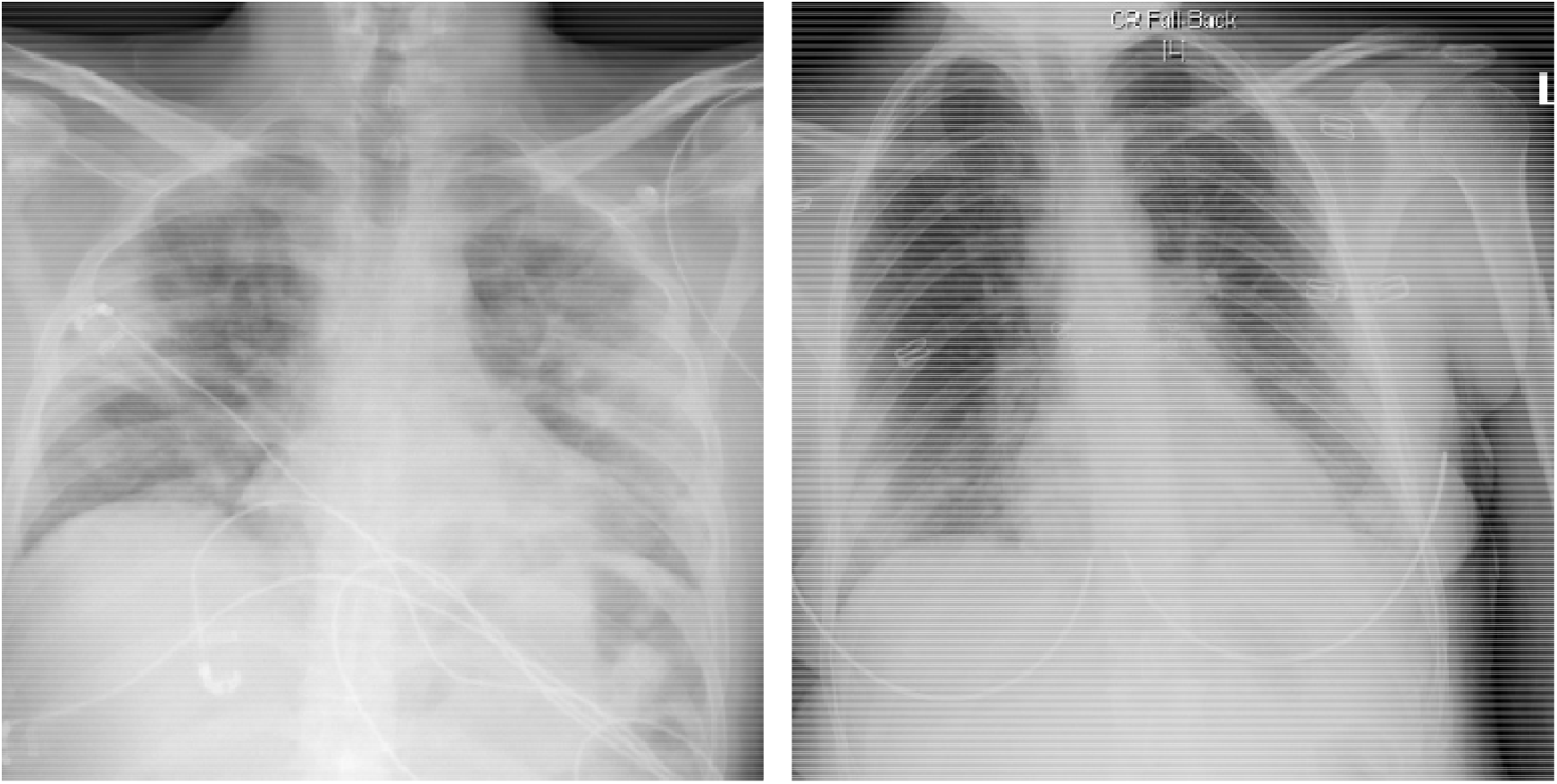
CXR’s of two different patients showing the most common abnormalities: bilateral, peripheral ground glass opacities and consolidation

**Table 3:** Radiological findings.

| Radiological findings (CXR) upon admission |  |  |  |  |  |
| --- | --- | --- | --- | --- | --- |
|  | All patients (n=150) | Mild cases (n= 105) | Moderate cases (n= 29) | Severe/Intensive care unit cases (n= 16) | P-value |
| **Predominant finding** |  |  |  |  |  |
| Normal | 72 (49.7) | 62 (60.2) | 7 (25.9) | 3 (20.0) | 0.000*** |
| Ground glass opacity | 42 (29.0) | 21 (20.4) | 13 (48.1) | 8 (53.3) | 0.002* |
| Consolidation | 26 (17.9) | 16 (15.5) | 6 (22.2) | 4 (26.7) | 0.488 |
| Linear atelectasis | 3 (2.1) | 3 (2.9) | 0 | 0 | 0.354 |
| Diffusion reticular | 1 (0.7) | 1 (1.0) | 0 | 0 | 0.795 |
| opacities |  |  |  |  |  |
| Reticulation | 1 (0.7) | 0 | 1 | 0 | 0.183 |
| Distribution within the lobe |  |  |  |  |  |
| Central | 10 (13.7) | 6 (14.6) | 2 (10.0) | 2 (16.7) | 0.833 |
| Diffuse | 21 (28.8) | 14 (34.1) | 4 (20.0) | 3 (25.0) | 0.494 |
| Peripheral | 42 (57.5) | 21 (51.2) | 14 (70.0) | 7 (58.3) | 0.378 |
| Distribution within the lung |  |  |  |  |  |
| Lower | 24 (32.9) | 12 (29.3) | 6 (30.0) | 6 (50.0) | 0.385 |
| Lower middle | 22 (30.1) | 10 (24.4) | 9 (45.0) | 3 (25.0) | 0.236 |
| Lower and middle and upper | 10 (13.7) | 8 (19.5) | 2 (10.0) | 0 | 0.089 |
| Diffuse | 10 (13.7) | 6 (14.6) | 2 (10.0) | 2 (16.7) | 0.833 |
| Peripheral | 2 (2.7) | 2 (4.9) | 0 | 0 | 0.309 |
| Middle | 2 (2.7) | 2 (4.9) | 0 | 0 | 0.309 |
| Upper | 1 (1.4) | 1 (2.4) | 0 | 0 | 0.559 |
| Upper and middle | 1 (1.4) | 0 | 0 | 1 (8.3) | 0.159 |
| No zonal predominance | 1 (1.4) | 0 | 1 (5.0) | 0 | 0.269 |
| Laterality |  |  |  |  |  |
| Bilateral | 53 (35.3) | 27 (25.7) | 16 (55.2) | 10 (62.5) | 0.000*** |
| Unilateral right | 12 (16.4) | 10 (24.4) | 2 (10.0) | 0 | 0.035* |
| Unilateral left | 8 (11.0) | 4 (9.8) | 2 (10.0) | 2 (16.7) | 0.805 |
| Progression |  |  |  |  |  |
| Stable | 62 (64.6) | 49 (74.2) | 11 (57.9) | 2 (18.2) | 0.001** |
| Worsen | 34 (35.4) | 17 (25.8) | 8 (42.1) | 9 (81.8) |  |

CXR: chest x-ray

### 3.3. Recovery

Patients stayed at the hospital for a mean duration of 9.2 days (SD: 3.9). Duration of stay in hospital ranged from two days to 23 days. At the end of the follow-up period a total of 94 patients (63.5%) recovered and 31.8% (n= 47) improved clinically but RT-PCR results were still positive. On the other hand, three patients (2.0%) did not fully recover and four patients (2.7%) deceased. The two patients who had mild cases died due to other reasons. There was no statistically significant difference based on the age regarding the recovery or whether the patient was symptomatic or asymptomatic upon presentation to hospital (p>0.05). The majority of the patients with mild cases improved or recovered, however, there was no statistically significant difference between cases from different severity and recovery rate (p>0.05) **(Figure 3)**.

**Figure 3:**
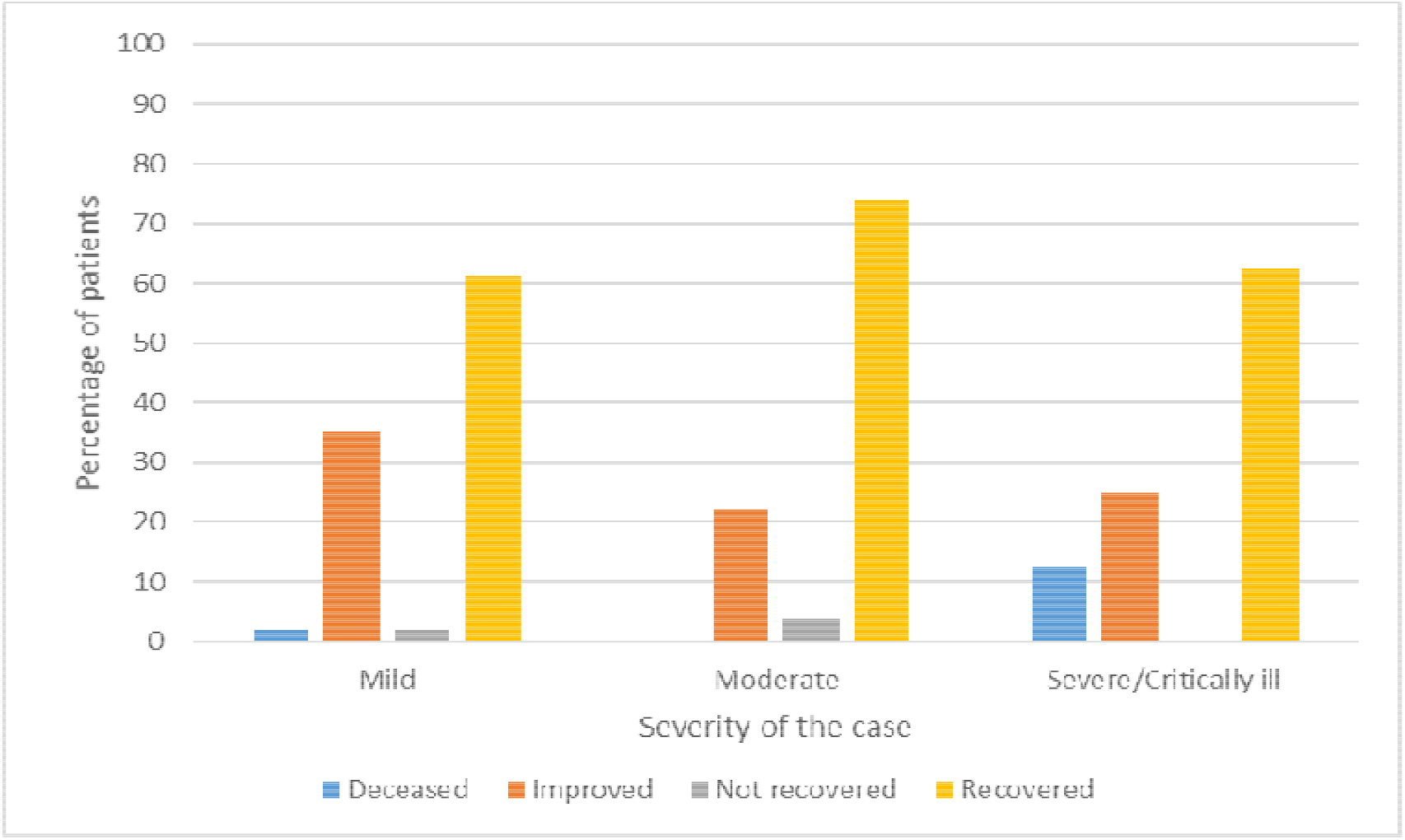
Recovery rates stratified by case severity

### 3.4. Therapeutic management

Beside supportive care, there were three main types of therapies that were prescribed to the patients for the management of COVID-19, this includes: a) antiviral therapy, b) antibiotics, and c) antimalarial medications **(Table 4)**.

**Table 4:** Initial treatment characteristics.

| Treatment therapy | Frequency (%) |
| --- | --- |
| Antiviral therapy |  |
| Combination of antiretroviral (lopinavir and ritonavir) and ribavirin | 14 (9.3) |
| Antimalarial therapy |  |
| Hydroxychloroquine | 25 (16.7) |
| Chloroquine | 15 (10.0) |
| Antibiotics therapy | 58 (38.7) |

## 4. DISCUSSION

To the best of our knowledge, this is the first and largest study to examine the clinical characteristics of COVID-19 in the Middle East region. We investigated clinical, radiological, and therapeutic characteristics of COVID-19 in a 150 hospitalised patients in Saudi Arabia. We found that around 89.0% of the cases were either mild or moderated and only 11.0% were either severe or critical. Our finding showed that the clinical severity of COVID-19 were of a milder presentation compared to results from China (12), Italy (13) and the United States (14, 15). These finding could be attributed to several factors including age and other demographics differences. The mean age in our study was 46.1 years (SD: 15.3) which was younger than the age reported in other studies. Several studies have reported poorer outcome among older population and patients with COVID-19 and comorbidities (16-18). However, it is difficult to draw a causal inference and we urge for further studies to investigate this association. In addition, it is important to highlight that the majority of the Saudi Arabian population are younger than 44 years (19).

Male patients with COVID-19 were more prevalent in our study compared to females, this was also similar to previous reports which highlight more males to be infected with COVID-19 (2, 14). These numbers could be because of men sex-based immunological differences or it could also be because of behavioural patterns such as smoking (20). In addition, comorbidities are more prevalent in men which could also be a reason for this difference (21), However, more researches must focus on gender differences and clinical outcomes with COVID-19.

Our study highlighted that around 28.8% and 26.0% of the study population had hypertension (HTN) and DM, these results were similar to previous reports that investigated the clinical characteristics of COVID-19 (1). Patients with DM and hypertension have an increased risk of complication of COVID-19 including acute respiratory distress syndrome (ARDS) (22), however, the mechanism of this remains un-investigated and it is unclear whether patients with uncontrolled blood pressure have a poorer outcomes of COVID-19 compared to patients with controlled blood pressure. In addition, Angiotensin-converting enzyme (ACE) inhibitors and angiotensin receptor blockers (ARBs) are two commonly prescribed medications for the management of HTN, and since SARS-CoV-2, binds to ACE2 in the lung, some theoretical theories have been raised about the benefits of these medications in the treatment of COVID-19 (23).

SARS-COV2 has been described to be similar to seasonal influenza, SARS-COV and MERS, this includes the fact that it is transmitted through respiratory droplets (24, 25). In addition, SARS-COV2 has similar symptoms to SARS-COV and MERS such as; fever, cough, and shortness of breath. This was reported in our study and it was also in line with previous studies (6, 13), however, SARS-COV2 has a higher case fatality rate in comparison to seasonal flu (0.1%) while it is also milder in comparison to other respiratory viruses such as SARS-COV (9.5%) and MERS (34.3%) (26). Besides this, COVID-19 is a highly infectious pathogenic (27, 28), with some reports suggested that half of the United Kingdom (UK) population has been infected without showing any symptoms or with having a mild course of the disease (29). Our study demonstrated that around 31.3% of the study sample were asymptomatic and had a mild disease. Mostly, these patients were identified through contact tracing and were isolated in the earlier course of the disease, whether this approach have any impact on the clinical course psychologically, this might need to be addressed in future studies. In addition, the majority of these patients had a contact with a confirmed COVID-19 patients which may raise concerns regarding the mechanism and the underlying inflammatory response in these patients. More researches are encouraged to investigate the characteristics of asymptomatic patients and if early detection and supportive treatment have a role in the clinical progression of the disease.

In our study, and unlike previous reports, nearly half of the patients presented with normal CXR, most of them were asymptomatic or had a mild disease. Furthermore, normal CXR at presentation may have a prognostic rule as only few numbers of those patients progressed into more severe cases. On the other hand, presence of ground glass opacity is linked with more aggressive course. The patterns found in abnormal exams were similar to the previously published reports and findings where peripheral, bilateral ground glass opacification (30).

Our study highlighted that around 26.7% of the patients received antimalarial treatment and around 9.0% received antiviral treatment, these medications have been suggested to have some beneficial effect to reduce the viral load and eliminate the disease, however, there are also uncertainties regarding their safety (31, 32). In addition, there has been debate about their efficacy in the treatment of COVID-19 with several trials are now in pipeline for the testing of these medications (33). To date, there is no treatment for COVID-19, and the main approach in the management of the disease is to provide supportive treatment and to control the symptoms including mechanical ventilator for critical cases (34). This study has some limitations. First, the number of patients included in the study were small. Second, the study population only included patients from a single-centre hospital in Saudi Arabia.

## 5. CONCLUSION

This case series provides clinical, radiological, and therapeutic characteristics of hospitalised patients with confirmed COVID-19 in Saudi Arabia. Our study demonstrates similar characteristics of COVID-19 to previously reported studies worldwide.

## Data Availability

The data that support the findings of this study are available from the corresponding author upon reasonable request.

## Conflict of interest

The authors have stated explicitly that there are no conflicts of interest in connection with this article.

## Authors contribution

Shabrawishi, Naser, Ghazawi and Alwafi had full access to all the data in the study and take responsibility for the integrity of the data and the accuracy of the data analysis. Shabrawishi and Alwafi had the original idea for this study. Shabrawishi, Naser and Alwafi contributed to the design of the study. Obaid and Alsharif contributed to the data collection. Naser and Alwafi contributed in the statistical analysis. Shabrawishi, Naser, Ekram and Alwafi wrote the first draft. All the authors contributed to interpretation and edited the draft report.

## Notes

### Competing Interest Statement

The authors have declared no competing interest.

### Funding Statement

No external funding was received.

